# Revealing the biology behind MRI signatures in high grade glioma

**DOI:** 10.1101/2023.12.08.23299733

**Authors:** Erika M Lewis, Lingchao Mao, Lujia Wang, Kristin R Swanson, Ramon F Barajas, Jing Li, Nhan L Tran, Leland S Hu, Christopher L Plaisier

**Affiliations:** School of Biological and Health Systems Engineering, Arizona State University, Tempe, AZ, 85287, USA; H. Milton Stewart School of Industrial and Systems Engineering, Georgia Institute of Technology, Atlanta, GA, 30332, USA; Mathematical Neuro-Oncology Lab, Department of Neurological Surgery, Mayo Clinic, Phoenix, AZ, 85054, USA; Department of Neurosurgery, Mayo Clinic, Phoenix, AZ, 85054, USA; Advanced Imaging Research Center, Oregon Health & Sciences University, USA; Department of Radiology, Neuroradiology Section, Oregon Health & Sciences University, USA; Knight Cancer Institute, Oregon Health & Sciences University, USA; Department of Cancer Biology, Mayo Clinic, Phoenix, AZ, 85054, USA; Department of Radiology, Mayo Clinic, Phoenix, AZ, 85054, USA; School of Computing, Informatics, and Decision Systems Engineering, Arizona State University, Tempe, AZ, 85281, USA

**Keywords:** MRI, tumor biology, immune infiltration, angiogenesis, image localized biopsies

## Abstract

Magnetic resonance imaging (MRI) measurements are routinely collected during the treatment of high-grade gliomas (HGGs) to characterize tumor boundaries and guide surgical tumor resection. Using spatially matched MRI and transcriptomics we discovered HGG tumor biology captured by MRI measurements. We strategically overlaid the spatially matched omics characterizations onto a pre-existing transcriptional map of glioblastoma multiforme (GBM) to enhance the robustness of our analyses. We discovered that T1+C measurements, designed to capture vasculature and blood brain barrier (BBB) breakdown and subsequent contrast extravasation, also indirectly reveal immune cell infiltration. The disruption of the vasculature and BBB within the tumor creates a permissive infiltrative environment that enables the transmigration of anti-inflammatory macrophages into tumors. These relationships were validated through histology and enrichment of genes associated with immune cell transmigration and proliferation. Additionally, T2-weighted (T2W) and mean diffusivity (MD) measurements were associated with angiogenesis and validated using histology and enrichment of genes involved in neovascularization. Furthermore, we establish an unbiased approach for identifying additional linkages between MRI measurements and tumor biology in future studies, particularly with the integration of novel MRI techniques. Lastly, we illustrated how noninvasive MRI can be used to map HGG biology spatially across a tumor, and this provides a platform to develop diagnostics, prognostics, or treatment efficacy biomarkers to improve patient outcomes.

## Introduction

Conventional magnetic resonance imaging (MRI) is a cornerstone for noninvasively diagnosing and devising treatment strategies for deadly high-grade glioma (HGG) tumors. Currently, MRI is primarily used to establish tumor boundaries that are readily observed as regions where the blood-brain-barrier (BBB) has been disrupted allowing for MRI contrast extravasation into the tumor interstitial space (*1*). Accurately delineating tumor boundaries is crucial for determining tumor burden, which translates clinically to the extent of surgical resection (*2*). The ubiquitous use of MRI in the clinical management of HGG tumors, coupled with the development of new MRI techniques, demands further study to elucidate the biological phenomena captured by imaging HGG tumors.

MRI measurements from HGG tumors have also been linked to tumor heterogeneity (*3, 4*), tumor percentage (*5*), cellular density (*6, 7*), and found to be predictive in prognostic and recurrence models (*8–10*). Simultaneous imaging and molecular characterization provide a platform for linking noninvasive biophysical imaging patterns to their molecular underpinnings. Examples include associations with EGFR mutational status (*11*) and CD49 immune cell presence (*12*) onto three-dimensional representations of a tumor. We hypothesize that additional insights into tumor biology can be extracted from MRI through integration of imaging and molecular measurements.

We conducted a system level integrative study to reveal the clinically relevant aspects of HGG tumor biology that can be captured through noninvasive MRI measurements. Establishing connections between MRI measurements and HGG tumor biology requires a spatially matched quantification of MRI alongside omics-scale molecular characterization. We generated a cohort of spatially matched MRI and transcriptomics characterized contrast enhancing (CE) HGG tumors (*13*). In the spatially matched cohort, we collected MRI measurements for T1-weighted contrast-enhanced (T1+C), T2-weighted (T2W), and two diffusion weighted imaging metrics: fractional anisotropy (FA), and mean diffusivity (MD). T1+C utilizes a contrast agent to identify areas of BBB disruption by registering where the contrast agent leaks out of vasculature into the surrounding tissue interstitia (*14*). T1+C is generally used as a surrogate for tumor presence and to define HGG tumor burden (*14*). T2W captures tissue water content with bright areas extending past the T1+C signal indicating vasogenic edema (*14*). An infiltrative tumor can extend into these regions of edema, and T2W is thus valuable alongside T1+C in guiding the extent of surgical resection (*14*). T1+C and T2W are commonly used to facilitate many aspects of HGG patient care, including initial diagnosis, treatment planning, and response assessment after first-line therapy. Advanced diffusion-based MRI techniques, such as FA and MD, have gained traction as essential complementary techniques in neuro-oncology. FA is a form of diffusion tensor imaging (DTI) that measures directionally dependent water diffusion (*14*). FA values measure perturbation from linear diffusion which is often used to characterize the risk for tumor infiltration of white matter tracts during pre-surgical mapping (*15*). MD is a form of diffusion-weighted imaging (DWI) that measures extracellular bulk water diffusion (*14*). MD has classically been used to measure water restriction related to cytotoxic edema and has been correlated with constrained diffusion due to cellular packing in neuro-oncology (*16*). Together, these MRI measurements quantify a wide range of biophysical phenomena that could potentially be used as noninvasive proxies for the underlying biology of HGG tumors.

Collecting spatially matched biopsies and MRI measurements from a tumor is a complex surgical undertaking, which leads to smaller cohort sizes. In this study, we boosted the signal in our analyses by overlaying the omics characterizations onto a previously constructed transcriptional map of glioblastoma multiforme (GBM). The gbmSYGNAL network was constructed using multi-omic profiles of 422 patient tumors from The Cancer Genome Atlas (TCGA) (*17*). The 5,194 genes in the gbmSYGNAL network are aggregated into 500 co-regulated gene expression signatures (biclusters). Biclusters contain genes that were linked to pathways and biological processes through functional enrichment analysis. We then linked enriched pathways and biological processes to the hallmarks of cancer (*17–20*). The hallmarks of cancer describe clinically relevant oncogenic functions acquired by all cancer types and are especially relevant to HGGs (*21*). We aim to identify MRI proxies for HGG tumor biology by discovering which spatially matched MRI measurements are predictive of clinically relevant tumor biology encapsulated within biclusters.

## Methods

### Patient recruitment and surgical biopsies

Patient recruitment took place at both Barrow Neurological Institute (BNI) and Mayo Clinic in Arizona (MCA) through IRB approved protocols at each institution. Informed consent was obtained prior to enrollment. Patients recruited for these studies underwent preoperative stereotactic MRI for surgical resection and had a histologically confirmed diagnosis of high-grade glioma (grade III or IV) and most of the patient tumors were IDH WT (25 IDH WT and 5 IDH mutant) (*13*). Most of the patient tumors were primary tumors (n = 23) and a smaller subset were recurrent tumors (n = 7). Characteristics of the patients, tumors and biopsies can be found in **Table S1**. The smallest possible diameter craniotomies were used to minimize brain shift. An average of 4-5 biopsies were collected from contrast-enhancing (CE) tumor regions separated by at least one cm using stereotactic surgical resection guided by conventional preoperative MRI (T1+C and T2W) (*13*). Biopsy locations were recorded via screen capture to allow co-registration of all MRI measurements.

### Conventional MRI to capture T1+C and T2W and general acquisition conditions

Patients were imaged 24 hours before stereotactic surgery using a 3T MRI scanner (Sigma HDx; GE-Healthcare Waukesha Milwaukee; Ingenia, Philips Healthcare, Best, Netherlands; Magnetome Skyra; Siemens Healthcare, Erlangen Germany). Conventional MRI included standard pre- and post-contrast T1-weighted (T1, T1+C, respectively) and pre-contrast T2-weighted (T2W) sequences as previously described (*13*). Briefly, T1 and T1+C images were acquired using inversion-recover prepped (IR-prepped) spoiled gradient recalled-echo (SPGR) (T1/TR/TE = 300/6.8/2.8 ms; matrix = 320 x 224; FOV = 26 cm; thickness = 2 mm). The gadolinium based contrast agent (GBCA) was gadobenate dimeglumine for patients recruited at BNI and gadobutrol for patients recruited at MCA. T2W images were acquired using fast-spin-echo (FSE) (TR/TE = 5133/78 ms; matrix = 320 x 192; FOV = 26 cm; thickness = 2 mm).

### DTI was used to capture MD and FA

DTI was performed using Spin-Echo Echo-planar imaging (EPI) [TR/TE 10,000/85.2 ms, matrix 256 × 256; FOV 30 cm, 3 mm slice, 30 directions, ASSET, B = 0,1000]. The DTI DICOM files were converted into NIfTI format using MRIConvert (http://lcni.uoregon.edu/downloads/mriconvert). DTI parametric maps were calculated using FSL (*22*) to generate whole-brain maps of MD and FA (*13, 23*).

### Image co-registration

Each MRI measurement was co-registered to the DTI B0 anatomical image volume using tools from ITK (*24*) and IB Suite (Imaging Biometrics, LLC) (*13*). This approach to co-registration minimizes potential distortion errors from resampling that can adversely affect DTI metrics. The co-registered data was resampled to an in-plane voxel resolution of ∼1.17 mm (256 x 256 matrix) and a slice thickness of 3 mm.

### Region of interest (ROI) generation and image feature extraction

The stereotactic location recorded for each biopsy during surgery defined a spatially matched MRI ROI measuring 8 x 8 x 1 voxels (9.6 x 9.6 x 3 mm). A board-certified neuroradiologist (L.S.H.) visually inspected all ROIs to ensure accuracy and that each biopsy specimen was in a CE region of the tumor (*13*). Biopsies near potential artifacts (e.g., the floor of the anterior skull base, middle cranial fossa superior to the mastoid air cells, or craniectomy plate/screws) or in locations expected to yield no tissue (e.g., resection cavity or central necrosis) were excluded from the study. The mean value was calculated from the voxels in each biopsy’s ROI for each MRI imaging technique, and these values were used for subsequent analyses linking MRI and transcriptomics.

### RNA-sequencing of image-localized biopsies

RNA was extracted from flash frozen tissue, libraries were constructed using the Illumina TruSeq v2 RNAseq kit, and paired-end reads were sequenced on an Illumina HiSeq 4000 sequencer. FASTQ files were aligned to the human transcriptome (GRCh38.p37) using STAR, read counts were compiled using htseq-count, and batch effect was corrected using ComBat-seq as previously described (*13*). Tumor purity was computed for all 75 samples with whole exome sequencing (WES) as previously described (*13*).

### Revising biclusters for use with RNA-seq data

The first principal component corrected for sign (eigengenes) for each gbmSYGNAL bicluster was computed from the 75 CE MRI samples with RNA-seq, tumor purity, and MRI measurements (*17, 25*). Mixed effects models (MEMs) from the statsmodels package (*26*) were used to associate each bicluster eigengene with the expression of each constituent gene across the 75 samples, using patient ID, tumor purity and IDH mutational status as random effects, and patient ID was used to account for repeated samples collected from the same patient tumor. Genes with a positive beta value (β > 0) and significant association (p-value ≤ 0.05) with the eigengene were included in the revised bicluster gene set definitions. Revised biclusters with at least five genes and an eigengene with a first principal component (PC1) variance explained ≥ 0.3 were included in subsequent analyses.

### Associating biclusters with MRI measurements

Revised bicluster eigengenes were tested for association with MRI measurements across the 75 samples using a MEM with patient ID, tumor purity, and IDH mutational status as random effects, and patient ID was used to account for repeated samples collected from the same patient tumor. Significant eigengene-MRI relationships were defined by a p-value ≤ 0.05.

### Linking MRI measurements to the hallmarks of cancer through gbmSYGNAL

In the construction of gbmSYGNAL, biclusters were associated with GO biological process (GO:BP) terms and then linked to the hallmarks of cancer using semantic similarity (*17*). Significant linkage to the hallmarks of cancer was defined as a significant MRI association with the bicluster eigengene and a Jiang-Conrath semantic similarity score ≥ 0.7 to a hallmark of cancer. For each MRI measurement, hypergeometric enrichment analysis was used to test the significance of the overlap between MRI and hallmarks of cancer associated biclusters (*17*). Significant enrichment was defined by a BH-adjusted p-value ≤ 0.05 and an overlap > 0.

### Linking MRI measurements to patient survival through gbmSYGNAL

In constructing gbmSYGNAL, biclusters were linked to patient survival using Cox proportional hazards regression with patient age as a covariate and replicated in at least one of three independent validation cohorts used in Plaisier et al., 2016 (*17*). This study defined a significant linkage between a bicluster and patient survival based on a p-value ≤ 0.05 and the hazard ratio in the same direction in all four cohorts (*17*). For each MRI measurement, hypergeometric enrichment analysis was used to test the significance of the enrichment between MRI and patient survival associated biclusters (*17*). Significant enrichment was defined by a BH-adjusted p-value ≤ 0.05 and an overlap > 0.

### Clinical and molecular data for the TCGA GBM cohort

The Pan-Cancer Atlas consortium contains standardized, normalized, batch-corrected, and platform-corrected multi-omics data for 11,080 participant tumors (*27*). This work used 422 complete multi-omic GBM patient tumor profiles to validate findings from MRI data. The TCGA GBM cohort microarrays were used for RNA expression (n = 422). TCGA aliquot barcodes flagged as “do not use” or excluded by pathology review from the Pan-Cancer Atlas Consortium were removed from the study. Overall survival (status and time to an event) for patients in the cohort were obtained from Liu et al. 2018 (*28*).

### Associating tumor infiltrating lymphocytes (TILs) to revised bicluster eigengenes in TCGA

Histologically determined quantification of TILs was available for 124 TCGA GBM tumors, where a value of 0 indicated TIL absence and values of 1 or 2 indicated TIL presence (*29*). For each revised bicluster eigengene, a t-test was performed comparing bicluster expression to TIL status (present vs. absent) across all 124 TCGA patients. Significant associations were determined by a BH-adjusted p-value ≤ 0.05. Hypergeometric enrichment analysis was used to test for enrichment between TIL and MRI associated biclusters, where significant enrichment was defined by a BH-adjusted p-value ≤ 0.05 and an overlap > 0.

### Deconvoluting immune cell abundance estimates

The CIBERSORTx deconvolution method was used to estimate immune cell abundance from bulk gene expression data (*30*). The MRI and TCGA cohorts were run through CIBERSORTx to estimate immune cell abundance using the LM22 signature matrix (*30*). Batch correction and quantile normalization were disabled, the relative run mode was selected, and 500 permutations were performed. Results were restricted to include T-cells (CD8+, CD4+ memory resting, and follicular helper), NK cells (resting and activated), and macrophages/microglia (non-activated, pro-inflammatory, and anti-inflammatory) to remain consistent with the infiltrating immune cell types previously identified in GBM tumors (*31–34*).

### Associating immune cell estimates to revised bicluster eigengenes

Deconvoluted immune cell fraction estimates were compared to bicluster eigengenes. Pearson correlations were used to test for associations between deconvoluted immune cell fractions for the TCGA cohort, because each patient corresponded to a single sample (*27*). Associations in the MRI cohort were computed using a MEM with patient ID, tumor purity, and IDH mutational status as random effects, where patient ID was used to account for repeated samples collected from the same patient tumor. Significant associations were identified for both the MRI and TCGA cohorts with a BH-adjusted p-value ≤ 0.05.

### Isolating hallmark-specific GOBP terms

For each MRI measurement, the genes from significantly associated revised biclusters were aggregated into two gene lists: one for positively associated revised biclusters, and a second for negatively associated revised biclusters. Genes were removed if they were members of two biclusters that had opposite beta value signs with the same MRI measurement (3 genes were removed in total). Functional enrichment analysis was performed on each gene list using the enrichr module from the gseapy package (*35*). Significant GO:BP terms were identified with a BH-adjusted p-value ≤ 0.05 and an overlap ratio of ≥ 0.1. Each GO:BP term was individually tested for significant semantic similarity with the hallmark(s) of cancer. Hallmark-specific GO:BP terms were identified with a Jiang-Conrath semantic similarity score ≥ 0.7. For each MRI measurement, GO:BP terms significantly associated with both the MRI measurement and the hallmark of cancer (‘Self sufficiency in growth signals,’ ‘Insensitivity to antigrowth signals,’ or ‘Tissue invasion and metastasis’) were isolated for further study.

### Associating vessel markup percentage to revised bicluster eigengenes in TCGA

Histologically determined percentages of vessel markup were available for 79 TCGA GBM tumors (*36*). A Pearson correlation was performed comparing each bicluster eigengene to vessel markup percentage. Significant associations were determined by a p-value ≤ 0.01. Hypergeometric enrichment analysis was used to test for enrichment between vessel and MRI associated biclusters, where significant enrichment was defined by a BH-adjusted p-value ≤ 0.05 and an overlap > 0.

### Calculating percentiles for MRI signal, bicluster expression, and gene expression

MRI measurements (T1+C), revised bicluster eigengenes (PITA-14, Targetscan-269, and Targetscan-541), and transcript levels (CD163) were normalized to scale between 0 and 1 across six MRI samples from patient BNI152 with RNA-seq, tumor purity, and MRI measurements.

## Results

### Cohorts of HGG tumors

We collected 75 spatially matched biopsies and MRI measurements from 30 HGG patient tumors (the MRI cohort, **Figure 1A, Table S1**). All 75 biopsies were characterized by RNA-seq and exome-seq to discover transcriptional signatures and tumor purity describing the underlying composition and molecular changes in each biopsy (*13*). An average of two and a half image localized biopsies (one to six biopsies) were collected from CE tumor regions separated by at least one cm, and average tumor purity, i.e., abundance of tumor cells in the biopsy, was 63% ± 20%. We accounted for the repeated sampling from the same tumor and tumor purity by incorporating them into our statistical tests.

**Figure 1.**
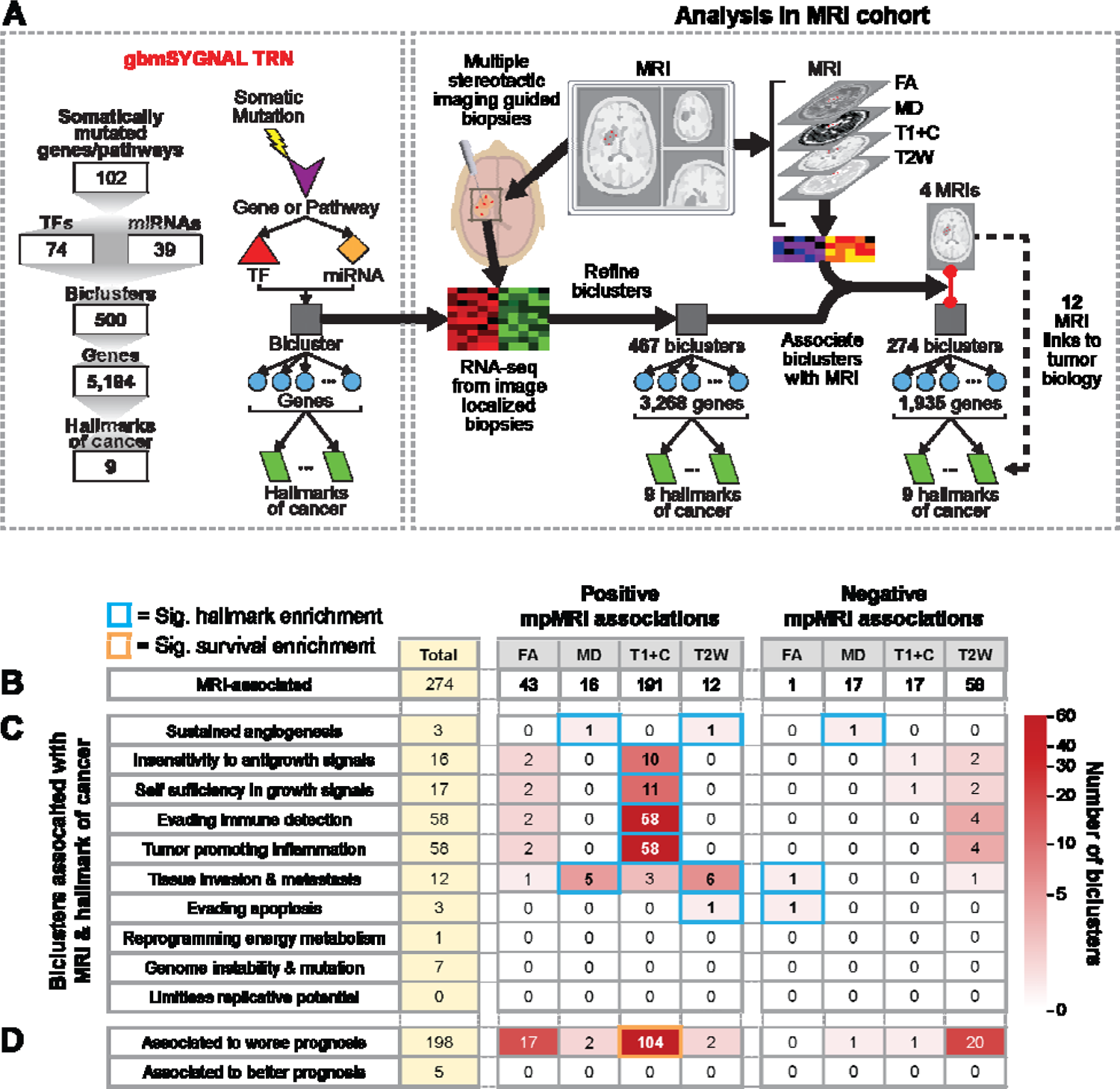
Associating MRI measurements with HGG tumor biology. (**A**) Biclusters from the gbmSYGNAL network were refined using RNA-seq data from the MRI cohort and were then used to associate MRI measurements and HGG tumor biology. (**B**) Number of biclusters associated with MRI measurements separated by positive and negative relationships with MRI signal. The Total column in yellow denotes number of biclusters associated with any MRI measurement, regardless of sign. (**C**) Number of biclusters associated with MRI and hallmarks of cancer separated by hallmark. Significant enrichment of MRI and hallmarks of cancer associated biclusters are denoted by a cyan outline. (**D**) Number of biclusters associated with MRI and patient survival separated by relationship with prognosis. Significant enrichment between MRI and patient survival associated biclusters are denoted by an orange outline.

The 30 tumors characterized in the MRI cohort cannot capture the full intertumoral heterogeneity observed between HGG tumors (*37*). We addressed this gap by incorporating the TCGA GBM cohort, which extensively profiled the transcriptomes of 422 patient tumors (*17, 27*). Previously we constructed the gbmSYGNAL network (*17*) to discover co-regulated gene expression signatures, or biclusters, that encapsulate the fundamental biological processes inherent to GBM tumors (i.e., hallmarks of cancer (*18, 19*)) (**Figure 1A**). We leverage the biology encapsulated by the 500 biclusters from gbmSYGNAL to enhance the depth and scope of our analyses with the MRI cohort. We also used the histological characterization of the TCGA GBM cohort to validate associations with immune cell infiltration (*29*) and angiogenesis (*36*). The integration of these two complementary cohorts culminates in a powerful platform for uncovering the HGG tumor biology captured by MRI measurements.

### Revising gbmSYGNAL biclusters for use with RNA-seq data

Technological differences between RNA-seq (MRI cohort) and microarray (TCGA cohort) are known to cause discrepancies in gene expression for some genes (*38, 39*). These discrepancies led to a lack of positive correlation for a minority of genes within the bicluster eigengenes of the MRI cohort (**Figure S1A**), which can impact downstream analyses. As a countermeasure, we devised a series of filters designed to eliminate genes exhibiting inconsistent expression patterns between platforms. This intervention was designed to ensure that the bicluster eigengene remains a robust representation of its constituent gene variance (**Figures S1A-D**). The implementation of the filters yielded a discernible impact: the exclusion of 1,926 genes (out of 5,194) and 33 biclusters (out of 500) from further analyses (**Table S2**). For the remaining 467 biclusters there was a reduction in the average number of genes per bicluster from 36 ± 11 to 25 ± 8. Importantly, the filters did not significantly alter the number of biclusters associated with each MRI measurement. The filters improved the results of downstream functional enrichment analyses (**Figures S1E-S1F**). In summary, the bicluster revision process removed genes marginally co-expressed in RNA-seq to improve bicluster coherence and bolstered the signal strength for subsequent functional enrichment analyses.

### Linking MRI measurements to the hallmarks of cancer

Our primary goal in these studies was to link MRI measurements to the underlying biology of HGG tumors. We first associated MRI measurements to revised bicluster eigengene expression using a MEM that corrects for tumor purity, repeated sampling from the same patient tumor, and IDH mutational status (MRI → bicluster; **Figure 1A**). Our data showed positive associations between all MRI measurements and the expression of at least 12 biclusters (**Figure 1B, Table S3**). Fewer negative associations were discovered that linked MRI measurements to the expression of biclusters (**Figure 1B, Table S3**).

Next, we integrated MRI associated biclusters to hallmarks of cancer using relationships previously defined in the gbmSYGNAL network (bicluster → hallmark(s) of cancer; **Figure 1A**) (*17*). Using the transitive property, we linked MRI measurements to the hallmarks of cancer with biclusters as the intermediaries (i.e., MRI → bicluster and bicluster → hallmarks of cancer, therefore MRI → hallmarks of cancer). The biclusters positively associated with MRI were significantly enriched with biclusters associated with seven hallmarks of cancer (BH-adjusted p-value < 0.05, **Figure 1C, Table S3**). The biclusters positively associated with T1+C were significantly enriched with biclusters associated with the immune-related hallmarks ‘Evading immune detection’ and ‘Tumor promoting inflammation,’ as well as the growth-related hallmarks ‘Insensitivity to antigrowth signals’ and ‘Self sufficiency in growth signals.’ Biclusters positively associated with MD and T2W were significantly enriched with ‘Sustained angiogenesis’ and ‘Tissue invasion & metastasis’ associated biclusters. Biclusters positively associated with T2W also had a significant enrichment with ‘Evading apoptosis’ hallmark associated biclusters. These results suggest that the MRI measurements (T1+C, MD, and T2W) might be used as noninvasive surrogates for seven hallmarks of cancer.

We also tested for enrichment between MRI and patient survival associated biclusters (**Figure 1D, Table S3**) (*17*). Biclusters positively associated with T1+C were significantly enriched with biclusters that predicted shorter patient survival (BH-adjusted p-value = 4.1 x 10^-13^). This observation is consistent with other studies that have recognized T1+C as a prognostic tool for HGG (*9, 40*).

### T1+C is associated with increased tumor infiltrating lymphocytes

The strongest association we observed in this study linked T1+C to biclusters enriched with the immune hallmarks of cancer (**Figure 1C**). We validated the association between T1+C and the immune hallmarks by testing whether the biclusters positively associated with T1+C were significantly enriched with biclusters associated with histologically-determined TILs in the TCGA cohort (*29*). We found that 151 of the 467 revised biclusters were significantly positively associated with TILs (T-statistic > 0; BH-adjusted p-value ≤ 0.05; **Table S4**). There was a highly significant enrichment between the TILs and T1+C associated biclusters (overlap = 125; BH-adjusted p-value = 1.1 x 10^-12^; **Figures 2A-D**). This striking overlap of TIL and T1+C associated biclusters supports that T1+C could be used as a proxy for tumor immune cell infiltration.

**Figure 2.**
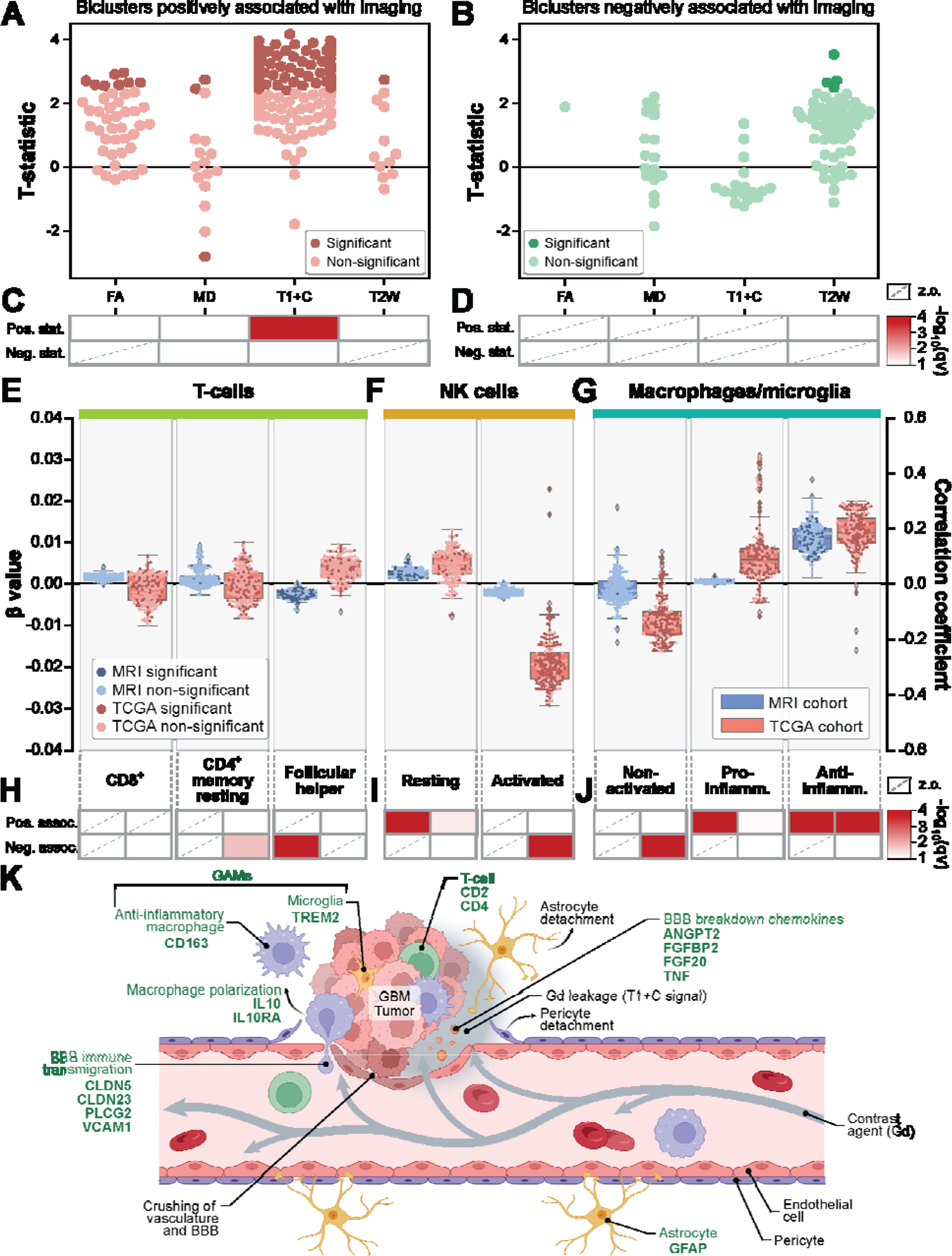
Validating the association of T1+C with infiltrating immune cells. Histologically determined TIL levels were tested for association with bicluster eigengene expression using a T-test for biclusters positively (**A**) and negatively (**B**) associated with MRI measurements. Significant associations with a bicluster are shown with darker red (**A**) or green (**B**) colors. Hypergeometric enrichment between TIL and positively (**C**) and negatively (**D**) MRI associated biclusters. “Pos. stat.” indicates a positive T-statistic and “Neg. stat.” indicates a negative T-statistic. Boxes with a dashed line indicate zero overlapping biclusters (z.o.). Associations between infiltrating immune cell estimates and T1+C bicluster eigengenes for T-cells (**E**), NK cells (**F**), and macrophages/microglia (**G**) using a MEM for the MRI cohort (left y-axis) and using correlation for the TCGA cohort (right y-axis). Significantly associated biclusters from the MRI cohort are denoted by darker blue and from the TCGA cohort are denoted by darker red. Hypergeometric enrichment between biclusters positively associated with T1+C and biclusters associated with T-cells (**H**), NK cells (**I**), and macrophages/microglia (**J**). “Pos. assoc.” indicates a positive association and “Neg. assoc.” indicates an inverse association. “Pro-inflamm.” indicates pro-inflammatory macrophages/microglia and “Anti-inflamm.” indicates anti-inflammatory macrophages/microglia. (**K**) Schematic of vasculature and BBB breakdown characteristic of HGG tumors. Green text represents the genes and functions included in T1+C biclusters.

### T1+C is associated with deconvoluted signatures of immune cell types

We then sought to identify the specific immune cell types infiltrating HGG tumors by examining associations between biclusters positively linked to T1+C and estimates of immune cell abundance. We focused our studies on the infiltrating immune cell types observed in HGG tumors (*31–34*), which included T-cells (CD8+, CD4+ memory resting, and follicular helper), NK cells (resting and activated), and macrophages/microglia (non-activated, pro-inflammatory, and anti-inflammatory). The choice of these cell types was confirmed by the observation that both T-cells and macrophages/microglia were the most abundant immune cell types in the estimates for the MRI and TCGA cohorts (**Figures S2A-C**).

Next, we tested for associations between the eigengenes from biclusters positively associated with T1+C and tumor-level estimates for the abundance of each immune subset (**Figures 2E-G**; **Table S5**). Associations between biclusters positively associated with T1+C and infiltrating immune cell estimates were considered significant if they replicated across the two independently collected MRI and TCGA cohorts. No consistent trends were observed for the T-cells between the two datasets (**Figures 2E** & **2H**). We did detect a significant positive relationship between biclusters positively associated with T1+C and resting NK cells (β-value and correlation coefficient < 0; BH-adjusted p-value ≤ 0.05; **Figures 2F** & **2I**). The most striking trend we observed was a strong positive relationship between biclusters positively associated with T1+C and anti-inflammatory macrophages/microglia (β-value and correlation coefficient > 0; BH-adjusted p-value ≤ 0.05, **Figures 2G** & **2J**). The increased infiltration of anti-inflammatory macrophages/microglia is strongly supported by literature reporting that these cells typically comprise a substantial fraction of HGG tumor bulk (*31–34*). A separate study also identified an increase of bone marrow-derived macrophages/microglia in the CE core of HGG tumors where T1+C signal is elevated, further supporting the link between T1+C and the presence of macrophages/microglia (*41*). Collectively, these findings provide strong evidence that elevated T1+C levels are linked to an increased presence of anti-inflammatory macrophages/microglia within the tumor microenvironment.

### Leaky vasculature and BBB breakdown leads to influx of contrast agent and immune cells

Our analyses showed evidence for a relationship between T1+C signal and tumor immune cell infiltration, but the underlying biophysical mechanism driving this association still needs to be explained. The biophysics of T1+C signal is based on the intravenous injection of a GBCA that is used for both enhancement of vasculature and to visualize HGG tumor burden (*14*). As HGG tumors grow, they crush the surrounding tissue and actively secrete enzymes that collectively lead to a loss in integrity of both the vasculature and BBB at various regions around the tumor (*42–46*). This leakiness allows GBCA out of the vasculature and across the BBB, where it generates a T1+C signal (**Figure 2K**). The genes from T1+C associated biclusters revealed the mechanisms for the breakdown of vasculature and BBB, and subsequent immune cell transmigration (**Figure 2K**). The biclusters included the angiogenic factors ANGPT2, FGFBP2, FGF20, and TNF that loosen the walls of vasculature and cell contacts of the BBB in brain metastases (*43, 46*). Physical crushing due to tumor growth in a confined space increases vasculature and BBB breakdown, allowing blood vessel contents to seep into the central nervous system (CNS) (*45*). The cell adhesion molecule VCAM1 mediates the adhesion of immune cells that, during extravasation, can lead to further loss of integrity for vasculature and the BBB (*47, 48*). These genes describe the mechanisms that disrupt the vasculature and BBB and allows GBCA to permeate into the tumor. These results corroborate the well-established mechanisms that generate the T1+C signal (*14*).

The biclusters positively associated with T1+C also contain genes describing immune cell infiltration (**Figure 2K**). The VCAM1 pathway members CLDN5, CLDDN23, and PLCG2 work with VCAM1 to signal immune cell transmigration (*48, 49*). Consistent with our deconvolution results, CD163 indicates the presence of anti-inflammatory macrophages that have been polarized by IL10 and IL10RA (*50, 51*) and TREM2 suggests that microglia are also present (*52*). Together, these macrophages and microglia constitute glioma-associated macrophages/microglia (GAMs). Additionally, markers CD2 and CD4 indicate that T-cells join anti-inflammatory macrophages as part of the invading immune cell population (*53, 54*). The co-occurrence of immune infiltration and T1+C signal in our results indicate that peripheral immune cells are likely crossing into the tumor at the same leaky sites that permit GBCA leakage (**Figure 2K**).

We observed that genes associated with vasculature and BBB breakdown and the transmigration of immune cells increase as T1+C signal increases. While the mechanism driving the elevation of T1+C signal due to vasculature and BBB breakdown is well-documented (*42–46*), it’s important to note that immune cell transmigration is a downstream consequence of vasculature and BBB breakdown. Meaning that T1+C does not directly quantify immune cell infiltration. Consequently, T1+C signal emerges as a valuable surrogate for vasculature and BBB breakdown and a meaningful albeit indirect indicator of immune cell infiltration within HGG tumors.

### T1+C is an indirect proxy for the proliferation of immune cells

Genes that increased when T1+C signal intensified were also associated with proliferation-related hallmarks of cancer (**Figure 1C**). The genes from the overlapping biclusters were strongly enriched with terms related to T-cell proliferation (**Figures 3A-3C**). This is consistent with the discovery of CD4^+^, CD8^+^, and follicular helper T-cell sub-population cell estimates in both the MRI and TCGA cohorts (**Figure S2A**). Other enriched pathways describe the process by which the T-cell antigen receptors begin the immune response through the tyrosine kinase signaling pathway (**Figure 3B**) to initiate the MAPK cascade and downstream ERK1/ERK2 and JNK pathways (**Figure 3C**) that result in proliferation (**Figures 3A-3B**) (*55–57*). The activation and proliferation of T-cells supports our original association between T1+C and infiltrating immune cells following vasculature and BBB breakdown.

**Figure 3.**
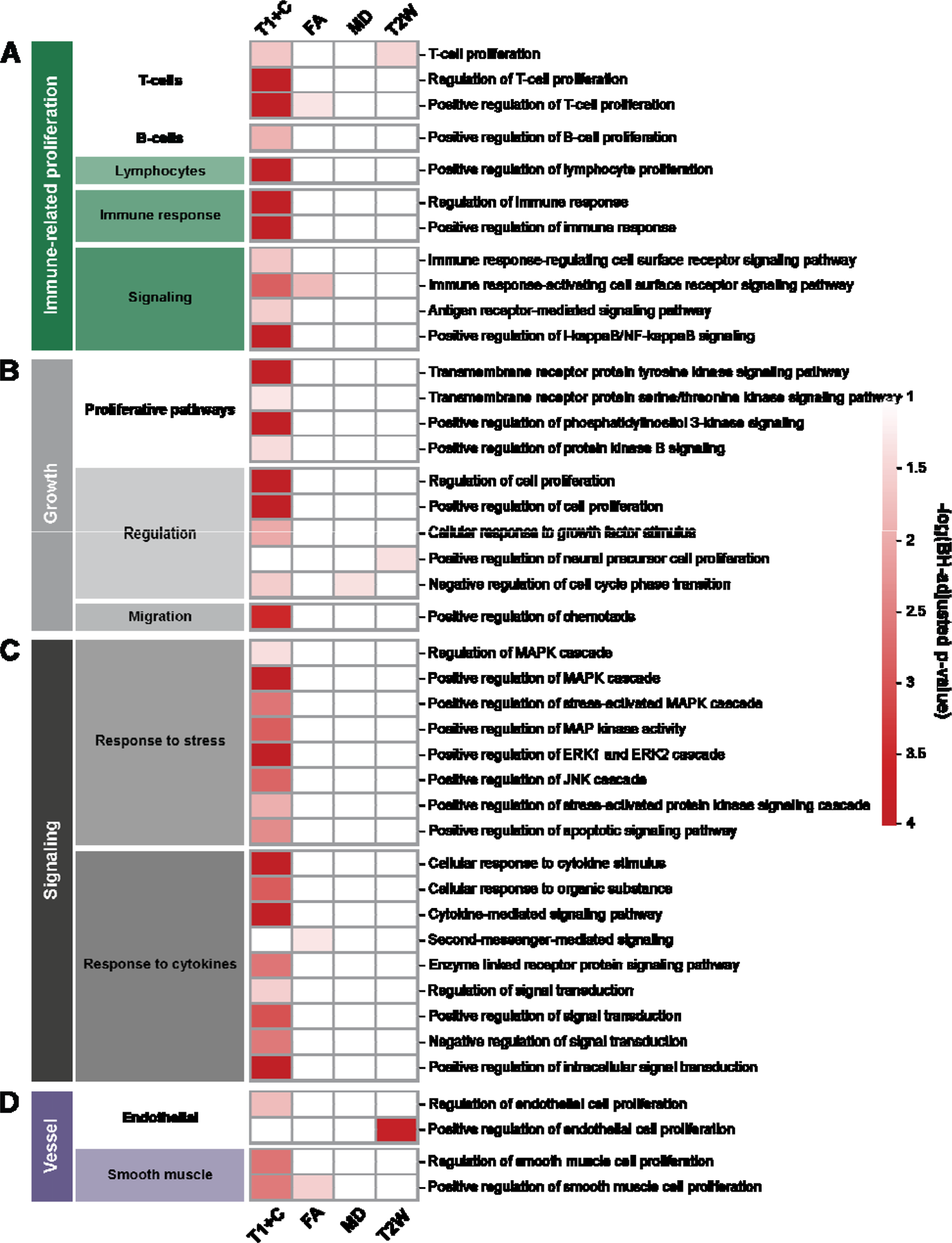
Proliferation-related biological processes enriched in imaging associated bicluster genes. Grouped by immune (**A**), growth (**B**), signaling (**C**), and vessel (**D**) related proliferation processes.

### T1+C is an indirect proxy for invading immune cell populations

Genes from the biclusters associated with ‘Tissue invasion & metastasis’ revealed a range of migration-related processes associated with increasing T1+C signal (**Figures 4A-4C**). These functions included immune cell migration, general migration processes, and coagulation (**Figures 4A-4C**). The immune-related migration functions split along distinct cell-type categories (**Figure 4A**), and the detection of monocytes (which include macrophages) and T-cell migration replicates our association between T1+C and immune infiltration (**Figure 2A-2J**). The migration-specific functions support that these immune cell populations are actively infiltrating HGGs.

**Figure 4.**
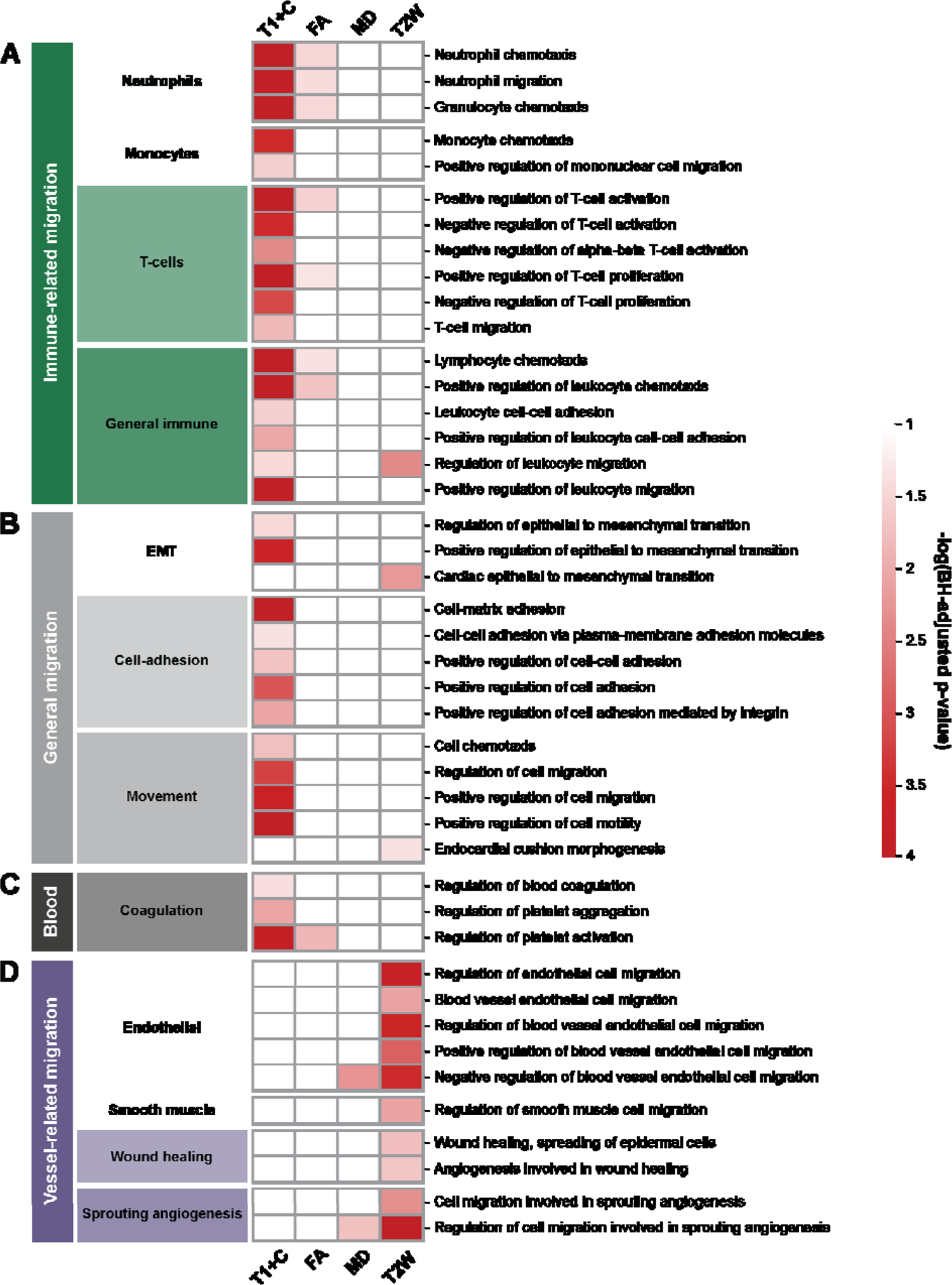
Invasion-related biological processes enriched in imaging associated bicluster genes. Grouped by immune (**A**), general (**B**), blood (**C**), and vessel (**D**) related migration processes.

### Using T1+C as a proxy for immune infiltration within a tumor

The heterogeneity of HGG tumors is a product of the varied anatomical constraints affecting each part of the tumor (e.g., proximity to vasculature, ventricles, skull, or white matter tracts), as well as the tumor microenvironment (e.g., overall cellular composition). We collected multiple samples from a tumor in the MRI cohort which allowed us to directly observe this heterogeneity at the molecular level through the RNA-seq and indirectly through our new noninvasive MRI proxies. The six biopsies from the tumor BNI152 are spatially arrayed throughout the CE parts of the tumor (**Figure 5A** & **B**). For each biopsy of patient BNI152’s tumor we show the average T1+C signal from the voxels containing the biopsy, the expression of three T1+C associated bicluster eigengenes, and the expression of a GAM marker gene CD163 (**Figure 5C** & **D**). The concordance between T1+C signal and the molecular data demonstrates how T1+C can be used to spatially map tumor immune cell infiltration (**Figure 5D** & **E**) that could be used to assist diagnosis, prognosis, or to assess the effect of immunotherapy on HGG tumors.

**Figure 5.**
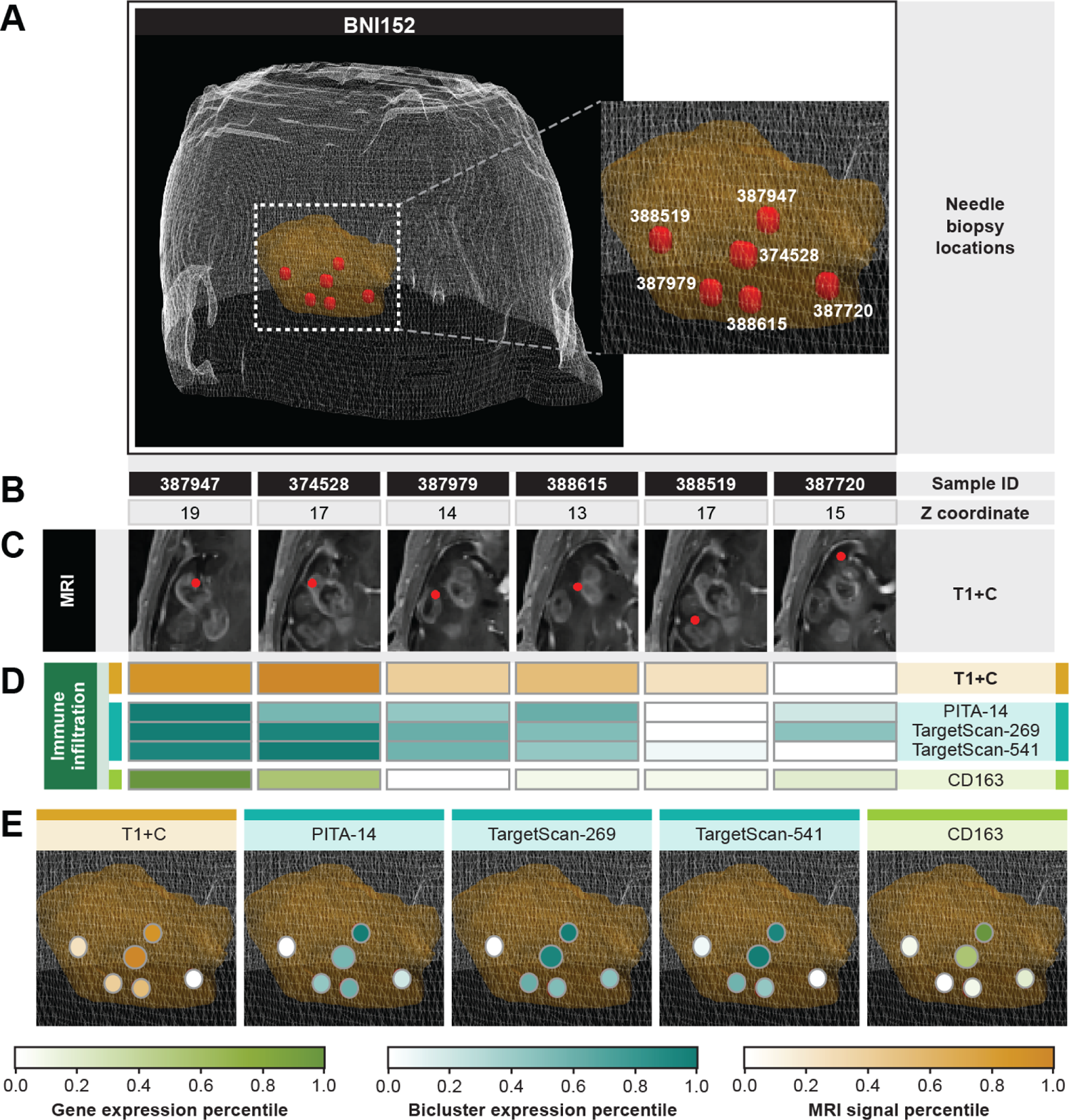
MRI captures immune infiltration across tumor biopsies. (**A**) Biopsy locations (red) within the tumor (yellow). (**B**) Z-coordinate of the slice of the MRI capture for each sample. (**C**) T1+C MRI image slice showing the biopsy location of each sample. (**D**) Percentiles of MRI signal, bicluster expression, and gene expression related to immune infiltration for each sample. (**E**) Percentiles of MRI signal, bicluster expression, and gene expression related to immune infiltration for each sample overlaid onto tumor spatial locations.

### Increased MD and T2W are associated with angiogenesis

Increased MD and T2W signal were associated with increased expression of a bicluster linked to the ‘Sustained angiogenesis’ cancer hallmark (**Figure 1C**), suggesting that MD and T2W may be proxies for angiogenesis. The low-strength magnetic field in clinical MRI adeptly captures the swift flow of fluids commonly present in vasculature. Consequently, both MD, which directly assesses water movement (*3, 58, 59*), and T2W, which signifies fluid presence (*14*), effectively capture the presence of vasculature, which is typically the origin of angiogenesis. We validated this relationship using a histologically determined measurement of angiogenesis (percent angiogenesis) from the TCGA cohort (*36*) (**Table S6**). First, we determined that 15 revised biclusters were significantly correlated with histologically determined percent angiogenesis (correlation coefficient > 0; p-value ≤ 0.01). Expression of four MD and four T2W associated biclusters were significantly correlated with percent angiogenesis which was more than expected by chance alone (**Figure 6A-D**; MD enrichment p-value = 1.2 x 10^-4^; and T2W enrichment p-value = 4.6 x 10^-5^). The relationship between angiogenesis and MD and T2W is reinforced by the significant overlap with histologically determined angiogenesis.

**Figure 6.**
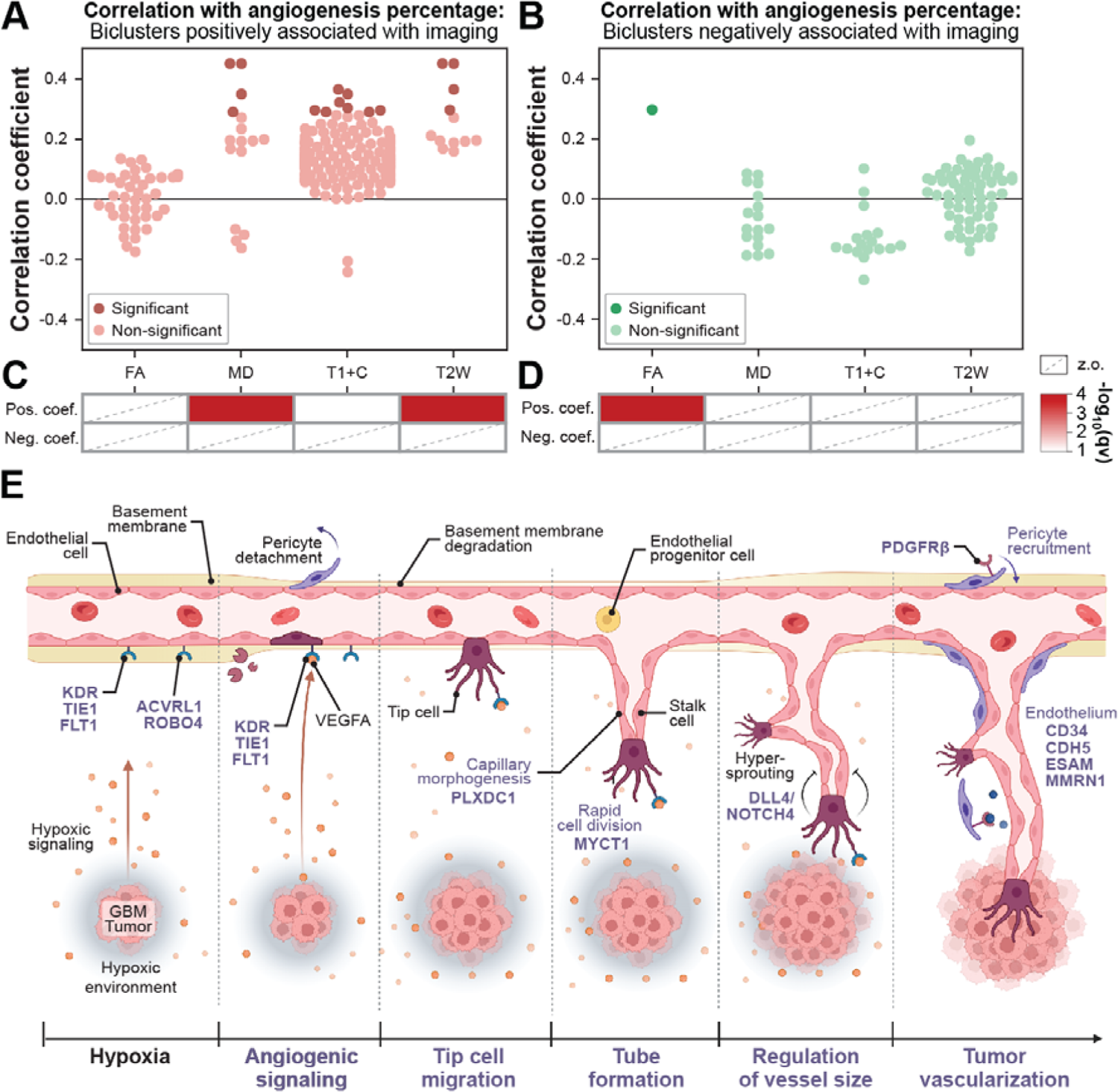
MD and T2W are associated with angiogenesis. Correlation between angiogenesis percentage and biclusters positively (**A**) and negatively (**B**) associated with MRI measurements. Hypergeometric enrichment between biclusters associated with angiogenesis percentage and biclusters positively (**C**) and negatively (**D**) associated with each MRI measurement. “Pos. coef.” indicates a positive correlation and “Neg. coef.” indicates an inverse correlation. Boxes with a dashed line indicate zero overlapping biclusters (z.o.). (**E**) MD and T2W biclusters contain genes (purple) from sprouting angiogenesis.

Further examination of the genes in the biclusters associated with MD and T2W revealed a wide range of biological processes that describe the progression of angiogenesis (**Figure 6E**). KDR, TIE1, and FLT1 are receptors for signals that initiate angiogenesis. ACVRL1 and ROBO4 are important receptors for the TGFβ and SLIT pathways, respectively, which promote cell growth and angiogenesis. PLXDC1 aids in capillary morphogenesis during the sprouting of new vessels. DLL4 and NOTCH4 regulate the growth of new vessels. PDGFRβ recruits pericytes to support the walls of new blood vessels. CD34, CDH5, ESAM, and MMRN1 indicate the presence of endothelial cells lining blood vessel walls. Finally, MYCT1 from the c-MYC pathway suggests a swift cell division process, ultimately contributing to the supply of cells necessary for vessel growth. In summary, there is significant histological and enriched angiogenic biology supporting MD and T2W being useful proxies for angiogenesis in HGG tumors.

### MD and T2W are proxies for cellular migration involved in angiogenesis

There are also positive associations linking MD and T2W to the biclusters associated with the ‘Tissue invasion & metastasis’ cancer hallmark (**Figure 1C**). The genes included in these ‘Tissue invasion & metastasis’ associated biclusters were enriched with angiogenesis related migratory processes (**Figure 4D**). The significantly enriched functions include the movement of endothelial and smooth muscle cells, which are the main cell types in the vasculature (**Figure 4D**). Cell migration involved in angiogenic wound healing and sprouting angiogenesis were also significantly enriched (**Figure 4D**). The formation of angiogenic sprouts requires the rearrangement and movement of endothelial and smooth muscle cells. These cell migratory functions are consistent with our findings that MD and T2W are linked to ‘Sustained angiogenesis’ and further support their value as proxies for angiogenesis in HGG tumors.

## Discussion

In these studies, we demonstrate that the systematic integration of image-localized biopsies with omics molecular profiles can reveal aspects of HGG tumor biology using noninvasive MRI measurements. Specifically, we provide evidence connecting T1+C signal to tumor immune infiltration and MD and T2W signals to angiogenesis. The T1+C MRI measurement directly captures the breakdown of vasculature and the BBB through the well-established leakage of contrast dye into the tumor. We also found that T1+C indirectly captures immune cell infiltration because the breakdown of vasculature and the BBB provides a permissive infiltrative environment. Moreover, our investigation revealed that MD and T2W effectively capture the intricate biological process of neovascularization. However, the molecular mechanism(s) responsible for generating the MRI signal in this context remains an area requiring further exploration. To fortify these connections between MRI measurements and tumor biology, we validated our findings using histology and by demonstrating the enrichment of well-established immune infiltration and angiogenesis pathways in associated gene expression signatures. Finally, we illustrated how noninvasive MRI can help understand HGG tumor biology spatially across a tumor, establishing a powerful tool for developing more accurate diagnostic, prognostic, and treatment efficacy biomarkers for HGG tumors.

Increased signal from T1+C indicated the presence of anti-inflammatory GAMs, and resting NK cells (*60*). These results are consistent with prior studies showing that HGG tumors are immunosuppressive (*61–63*), and this trait is partially attributed to anti-inflammatory GAMs (*61–63*). Future studies with spatially matched cohorts that unbiasedly quantify all immune cell types from HGG tumors directly using single-cell and spatial transcriptomics techniques will be able to better characterize the relative contributions of all immune cell types in areas of elevated T1+C signal.

Recent studies have demonstrated the potential of MRI biomarkers for angiogenesis by linking survival benefits from anti-VEGF therapy to the apparent diffusion coefficient (ADC) (*64, 65*), which quantifies the same biophysical phenomenon as MD. Patients whose tumors had higher ADC levels showed nearly double the overall survival when treated with anti-VEGF compared to those with lower ADC levels. The MRI measurements from the anti-VEGF studies and our MRI cohort were taken prior to therapy. Our research offers evidence that ADC effectively quantifies angiogenesis, explaining its utility as a biomarker for predicting the response to anti-angiogenic therapy. This example showcases how our approach of linking MRI measurements to tumor biology can be used to inform clinical studies to develop noninvasive biomarkers.

T1+C and T2W are commonly used to facilitate many aspects of HGG patient care, including initial diagnosis, treatment planning, and response assessment after first-line therapy. Our results enhance the utility of these commonly used MRI measurements by showing they predict HGG tumor biology. Our discovery that noninvasive MRI measurements are proxies for immune cell infiltration and angiogenesis lay the groundwork for capturing more HGG tumor biology. Future studies will incorporate new MRI measurement techniques and test their ability to capture novel aspects of HGG tumor biology. The long-term goal of this work will allow clinicians to noninvasively monitor HGG tumor biology, allowing them to improve diagnosis and prognosis, and respond to treatment in real time, so they can adjust treatment and improve patient survival.

## Data Availability

All data produced in the present study are available upon reasonable request to the authors.

https://www.synapse.org/#!Synapse:syn52256644

https://gdc-hub.s3.us-east-1.amazonaws.com/download/TCGA-GBM.htseq_counts.tsv.gz

https://github.com/HuLiLab/Multi-Regional-GBM-Imaging-and-Genetics

## Acknowledgments

This work was supported by NIH-NINDS Award # 1R01NS123038-01, and 1R01NS119650-01. The authors acknowledge the Glioma Biology Protocol Team for their work in collecting the MRI cohort and the Cancer Genome Atlas Research Network for the TCGA Pan-Cancer Atlas multi-omic patient tumor profiles. We also thank Dr. Benjamin Bartelle and Dr. Scott Beeman for sharing their knowledge of MRI. Figures 2K and 5E were created with BioRender.com.

## Author contributions

K.R.S., R.F.B., J.L., N.L.T, L.H., and C.L.P. conceived the project. K.R.S, N.L.T, and L.H. provided resources through the MRI cohort data. C.L.P. provided resources through the TCGA cohort data. E.M.L., L.H. and C.L.P curated data from both studies. E.M.L. and C.L.P. conducted formal analysis, investigations, and validations. E.M.L., L.M., L.W., J.L., and C.L.P. contributed to visualizations. E.M.L., L.H., and C.L.P. wrote the manuscript. All authors contributed to the review and editing of the manuscript. N.T.L., L.H., and C.L.P. contributed to supervision and project administration. K.R.S., R.F.B., J.L., N.L.T, L.H., and C.L.P. contributed to the funding to support this project.

## Notes

### Competing Interest Statement

The authors have declared no competing interest.

### Author Declarations

Patient recruitment took place at both Barrow Neurological Institute (BNI) and Mayo Clinic in Arizona (MCA) through IRB approved protocols at each institution. Informed consent was obtained prior to enrollment. All datasets analyzed in the current study, including whole exome sequencing and RNA sequencing, are publicly accessible, with the Synapse (https://www.synapse.org/#!Synapse:syn52256644). The publicly available bulk RNA-seq data for TCGA-GBM were obtained from the UCSC Xena browser [https://gdc-hub.s3.us-east-1.amazonaws.com/download/TCGA-GBM.htseq_counts.tsv.gz]. The input data for the imaging analysis can be found at https://github.com/HuLiLab/Multi-Regional-GBM-Imaging-and-Genetics Source data are provided.

