## Supplemental Information for "Revealing the biology behind MRI signatures in high grade glioma"

### **Table of contents**

### **Supplementary figures**

**Figure S1** Filtering pipeline to revise gbmSYGNAL biclusters for use with RNA-seq data.

**Figure S2** Association between infiltrating immune cells and mpMRI is lost when all mpMRI biclusters are considered.

### **Supplementary tables**

**Table S1.** Patient characteristics.

**Table S2.** Revised bicluster definitions.

**Table S3.** Summary of results linking mpMRI measurements, bicluster eigengenes, the hallmarks of cancer, and patient survival.

**Table S4.** Summary of t-tests relating bicluster eigengenes and tumor infiltrating lymphocytes in the TCGA cohort.

**Table S5.** Summary of associations relating bicluster eigengenes and immune cell fractions in the mpMRI and TCGA cohorts.

**Table S6.** Summary of Pearson correlations between bicluster eigengenes and vasculature percentage in the TCGA cohort.

### **Supplemental Figures**

**Figure S1.** Filtering pipeline to revise gbmSYGNAL biclusters for use with RNA-seq data. The β coefficients from the MEM relating gene expression to its parent bicluster eigengene, before (**A**) and after (**B**) bicluster revision. Each dot represents a gene, and the results displayed are from the biclusters positively associated with the MD mpMRI measurement to provide an example of revision. The revision pipeline removed genes with either opposite (β < 0) or non-significant (p-value ≤ 0.05) expression patterns to their parent bicluster. (**C**) Number of genes in each bicluster before (left) and after (right) being revised. Each dot represents a bicluster. Any revised biclusters with less than 5 genes, below gray dashed line, were dropped from further analyses. (**D**) Variance explained of PC1 at each level of filtering, where each dot represents a bicluster eigengene. (**E**) Number of biclusters significantly associated (p-value ≤ 0.05) to each mpMRI measurement. Solid lines indicate positive (β > 0) associations and dashed lines indicate negative (β < 0) associations. (**F**) Number of GOBP terms significantly enriched (BH-adjusted p-value ≤ 0.05) from the genes of the biclusters significantly associated (p-value ≤ 0.05) to each mpMRI measurement at each filtering level. Var. exp. = variance explained; pos. assoc. = positive association (β > 0); sig. assoc. = significant association (BH-adjusted p-value ≤ 0.05); pos. and sig. assoc. = positive and significant association (β > 0 and BH-adjusted p-value ≤ 0.05).

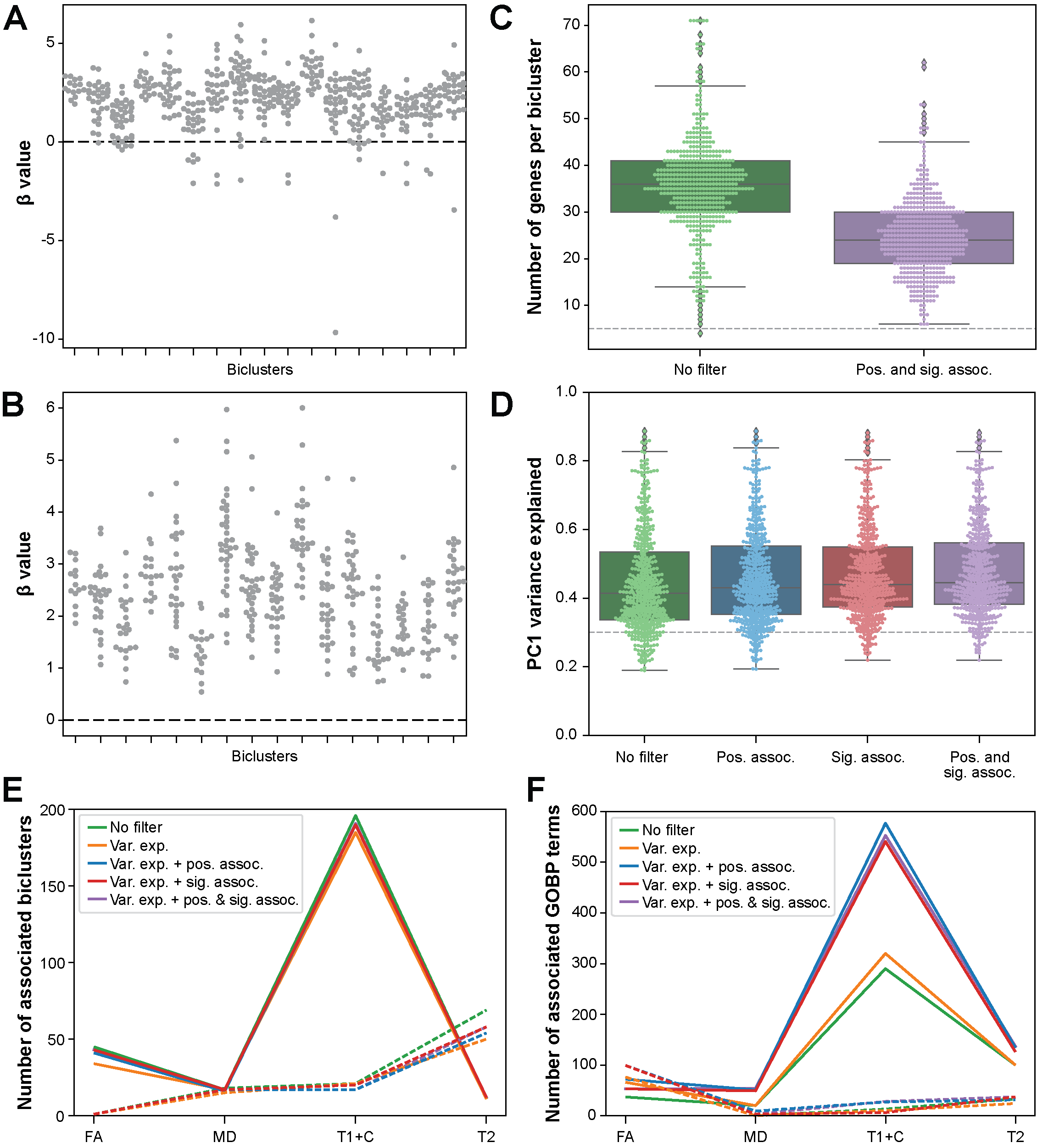

**Figure S2.** Infiltrating immune cell populations are present in HGG tumors. Deconvoluted infiltrating immune cell fractions in mpMRI and TCGA for: (**A**) T-cells, (**B**) NK cells, (**C**) macrophages/microglia, (**D**) B-cells, (**E**) DCs, (**F**) mast cells, (**G**) and granulocytes. Rest. = resting state, Act. = activated state, Follic. Helper = follicular helper T-cells, Mono = monocytes, Non-act. = non-activated macrophages/microglia, Pro-inflamm. = pro-inflammatory macrophages/microglia, Anti-inflamm. = anti-inflammatory macrophages/microglia, Mem. = memory B-cells, Eosin. = eosinophils, and Neutr. = neutrophils.

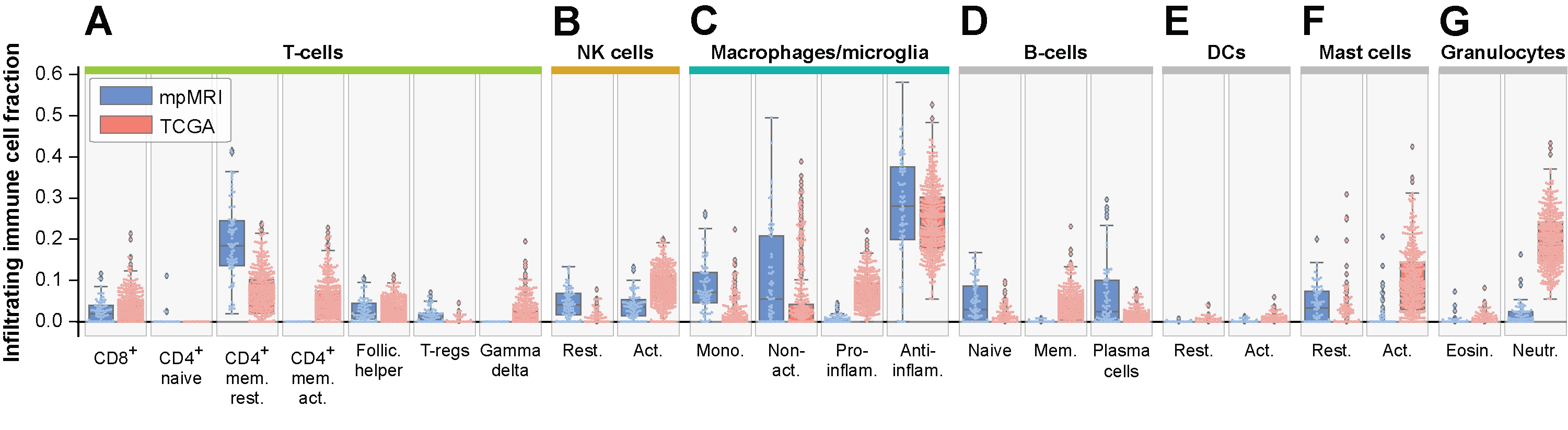

**Figure S3**. Anti-proliferative biological processes enriched in imaging associated bicluster genes. Grouped by immune (**A**), growth (**B**), signaling (**C**), and vessel (**D**) related anti-proliferation processes. Colors inside the box indicate the significance of the association between the GOBP term and the genes associated with the mpMRI measurement.

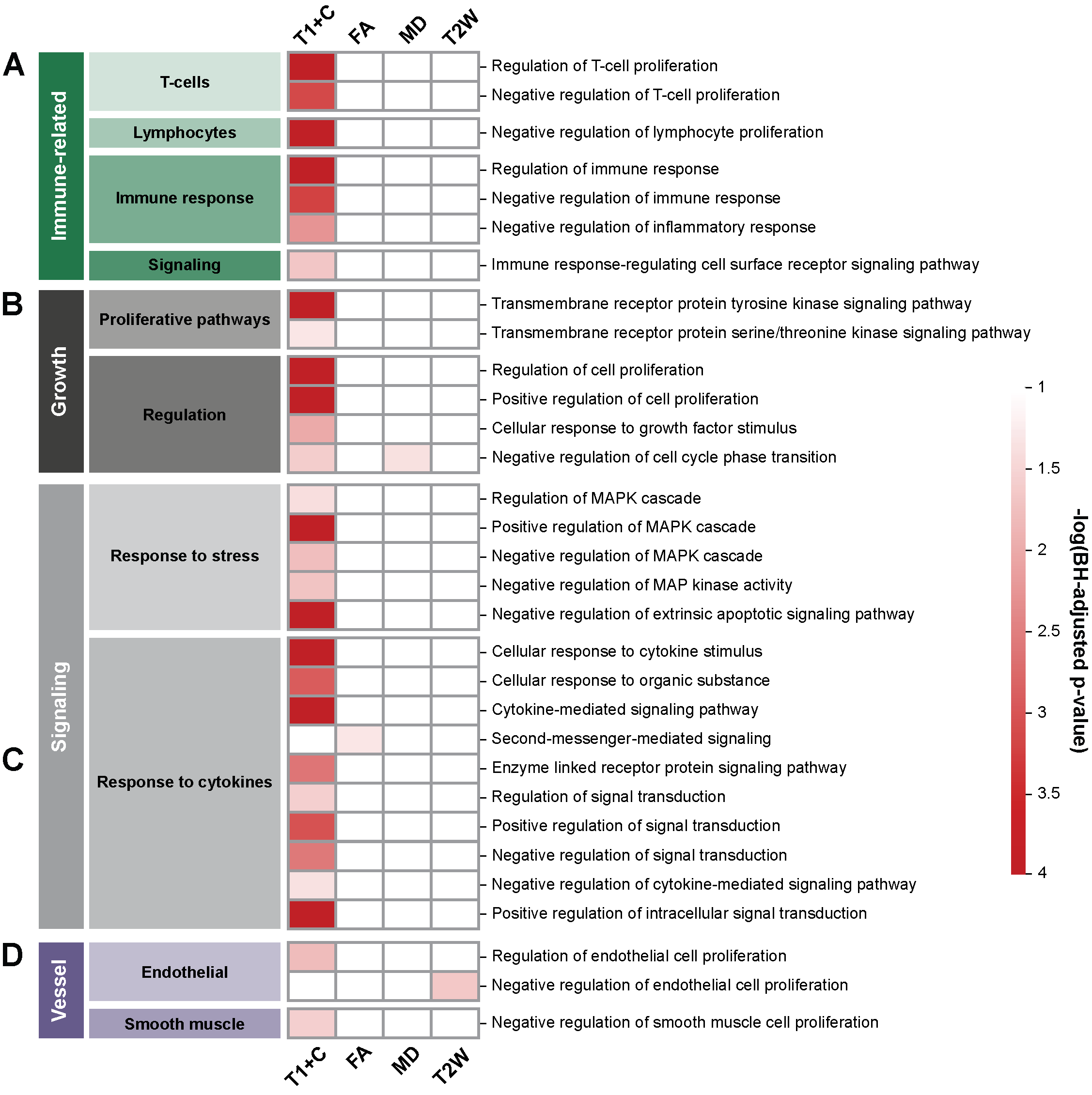
